# Exploring the Impact of Non-Emergency Inter-Hospital Neonatal Transport of Preterm Infants: Study Protocol for a Scoping Review

**DOI:** 10.1101/2025.03.19.25322589

**Authors:** L.A. Derks, M.L.J. Partouns, M.R. Koolen-De Konink, A.J. Apperloo, J.H.L. Wagenaar, A.M. Tjiam, H.R. Taal

## Abstract

**Background:** Within a regionalized system for neonatal intensive care, non-emergency transfers away from tertiary NICU hospitals occur frequently, for example due to a stabilized condition of the child, due to capacity issues, or for semi-elective surgical treatment. Although the negative impact of transport of critically ill preterm neonates has been extensively described, the effects of non-emergency transport specifically is not widely reported on. This scoping review aims to examine the current literature on how non-emergency neonatal inter-hospital transfers of preterm infants affect both the child and their parents.

**Methods:** A scoping review will be conducted following the PRISMA-ScR guidelines. Medline, Embase and CINAHL data will be searched for English or Dutch language empirical research publications or guidelines. Excluded are (systematic) reviews, (educational) simulation-based research, commentary, protocols and conference abstracts. Screening, data extraction and quality assessment will be performed by at least two independent authors. Quantitative and qualitative data will be assessed separately. Outcome measures include any information of the clinical situation/stability of the child before, during and/or after transport, and any parent reported outcome and experience measures regarding transfer experiences.

## Introduction

### Background and Rationale

Preterm birth is defined by the World Health Organization (WHO) as live birth before completing 37 weeks of pregnancy. Preterm births can be further subdivided into moderate of late preterm (32-37 weeks), very preterm (28-<32 weeks) or extremely preterm (<28 weeks). (1) In the Netherlands, there are approximately 10.000 live preterm births (7% out of a total of 160.000 live births) each year. Children born prematurely require a high level of care: on average, preterm infants born before 32 weeks in the Netherlands spend 17 days in a tertiary level Neonatal Intensive Care Unit (NICU). (2, 3) Neonatal intensive care services are provided by nine tertiary referral hospitals spread across the country. Additionally, surrounding regional hospitals provide lower levels of neonatal care in neonatal post-IC/high care or medium care units. This organization of care could be described as “regionalized” or “network-based”, and is common across many developed countries. (4-11) A regionalized neonatal care structure allows for high quality, highly specialized neonatal care to be delivered to extremely fragile preterm infants, (4, 12) and simultaneously allows for non-emergency transfers from the tertiary NICU to a hospital closer to home once the infant reaches a stable condition. Apart from these transfers to lower-level care settings, non-emergency transfers also occur for other reasons, for example, if there is a capacity problem, or when the infant develops a specific condition that needs semi-elective treatment that can only be performed elsewhere. In the Netherlands specifically, neonatal pediatric surgery is centralized (13): six out of nine tertiary NICU hospitals have a pediatric surgical unit. Moreover, each pediatric surgical unit is only specialized in a few specific indications, for example: only two out of six centers provide the surgical management of congenital diaphragm hernia’s (CDH). (13) In addition to neonatal pediatric surgery, treatment of retinopathy of prematurity with laser coagulation is only performed in five out of nine tertiary NICU hospitals. Although non-emergency transfers (i.e. transfers to lower care settings, transfers for surgery, or any other non-emergency inter-hospital transport) are regularly required given the current organization of neonatal (intensive) care in the Netherlands, they are generally considered to be less than ideal, not only because of costs and logistical challenges, but also because of possible negative effects on the well-being of the child and its parents. Due to the non-urgent nature of these transfers, health care professionals should carefully weigh possible upsides and downsides of transporting these infants within the broader context of the organization of neonatal care. Moreover, in the case of semi-elective surgery, the alternative of having the surgeon come to the child instead of transferring the child should also play a role within the decision-making process.

### Objective(s) and research questions

With this scoping review, we want to examine the current literature on how non-emergency neonatal inter-hospital transfers of preterm infants affect both the child and their parents. The main research questions are:

- What is the impact of non-emergency neonatal inter-hospital transport on the health or physical condition of the child?
- What are the effects of non-emergency neonatal inter-hospital transport on the well-being of parents?

To answer these research questions, we also drafted the following explorative questions:

- What are commonly used outcome measures to assess the effects of transport on the health or physical condition of the child?
- What are commonly used outcome measures to assess the effects of transport on the well-being of parents?

## Methods

This protocol was developed using the JBI Manual for Evidence Synthesis (14) and the extension of the PRISMA statement (PRISMA-ScR) (15). The PRISMA-ScR checklist was filled out to the extent possible and can be found as a supplemental file to this protocol (Supplemental 1). This protocol was registered under https://doi.org/10.17605/OSF.IO/Q5YR9 in Open Science Framework (OSF).

### 1.1 Eligibility Criteria

Eligibility criteria for this scoping review are defined using the Participants Concept Context (PCC) framework. (16) An overview of eligibility criteria is shown in Table 1.

**Table 1:** PCC Framework.

| Frame | Eligibility criteria |
| --- | --- |
| P (Participants) | Preterm infants born at < 37 weeks gestational age. |
| C (Concept) | INCLUSION: Non-emergency medical transport of clinically stable preterm infants, including transfers to lower care settings, transfers for semi-elective surgery, or any other non-emergency inter-hospital transport.<br>EXCLUSION:<br>• Antenatal, maternal or intra-uterine/partum transfers<br>• Acute-stage transfers<br>• Readmissions after discharge home<br>• Transfers within the first three days of life |
| C (Context) | Patient or parent-centered empirical research publications that published before 21 October 2024 in Dutch or English. |

#### 1.1.1 Participants

The target population of this scoping review are preterm infants (as defined by the WHO: born < 37 weeks gestational age). Research publications that only report on medical transport of full-term infants will be excluded.

#### 1.1.2 Concept

This scoping review is centered around the impact of non-emergency medical transfers of clinically stable preterm infants. This excludes in-utero transport, i.e. transport of the mother before or during childbirth, or readmissions after initial discharge home. Acute-stage transfers of clinically unstable infants that require intensification of treatment or life support at another hospital fall outside the scope of this review. Publications that only report on transfers that take place within the first three days of life will therefore also be excluded.

#### 1.1.3 Context

Our research questions should be placed in the context of a regionalized or centralized neonatal care system. Only Dutch or English language empirical research publications that involve patients (or their parents) published before October 21^st^, 2024, are eligible for inclusion. Excluded are (systematic) reviews, (educational) simulation-based research, commentary, protocols and conference abstracts.

### 1.2 Information Sources and Search Strategy

Information sources for this scoping review can be original research publications or guidelines. For original research publications, different study design will be considered for inclusion, including, but not limited to, randomized or non-randomized controlled trials, prospective and retrospective cohort studies, case-control studies, cross-sectional studies and qualitative studies. Excluded are (systematic) reviews, (educational) simulation-based research, commentary, protocols and conference abstracts.

In an initial search of Medline (PubMed) and Embase, relevant text terms, index terms and definitions were identified. A limited search with identified terms was performed and was used to find any relevant additional terms or sources. Subsequently, a full search strategy was developed with the help of an expert from the Erasmus MC Medical Library. The search terms can be found in Supplemental 2. Databases that will be searched are Medline, Embase and CINAHL. The search is limited to articles in Dutch or English only. If this scoping review is not finalized within six months after the last day searched, the Medical Library will update the search.

If the authors come across any other relevant papers, guidelines, websites or grey literature among the cited sources of included articles, those references will be reviewed by the authors for possible inclusion. Dutch or English language databases of national or regional guidelines will be searched for any relevant sources on neonatal transport.

### 1.3 Study Selection

Planned study selection procedures are summarized in Figure 1. Publications identified using the full search strategy will be exported into EndNote X9 and duplicates will be removed. The deduplicated RIS file will be imported into Covidence. Three authors (LAD, MLJP and MRK) will independently screen titles and abstracts to determine potential eligibility. Prior to screening, the use of Covidence will be piloted by five authors (LAD, MLJP, MRK, AJA, JHLW) with a random sample of 10% of the publications found, so that the screening process can be refined if needed. During abstract and title screening, two unanimous votes are needed for a decision. Conflicts will be resolved by discussion with the research team. Full-text retrieval will be facilitated by Covidence, or if necessary, articles will be manually retrieved using EndNote in RIS or BibTeX format. Full-text examination of publications selected during initial screening will be performed by the same three authors. The decision to include or exclude publications should be completely unanimous; any disagreements are resolved during a consensus meeting in the presence of two additional authors (AJA & JHLW). Reasons for exclusion of publications during full-text examination will be recorded in Covidence and reported in the scoping review.

**Figure 1:**
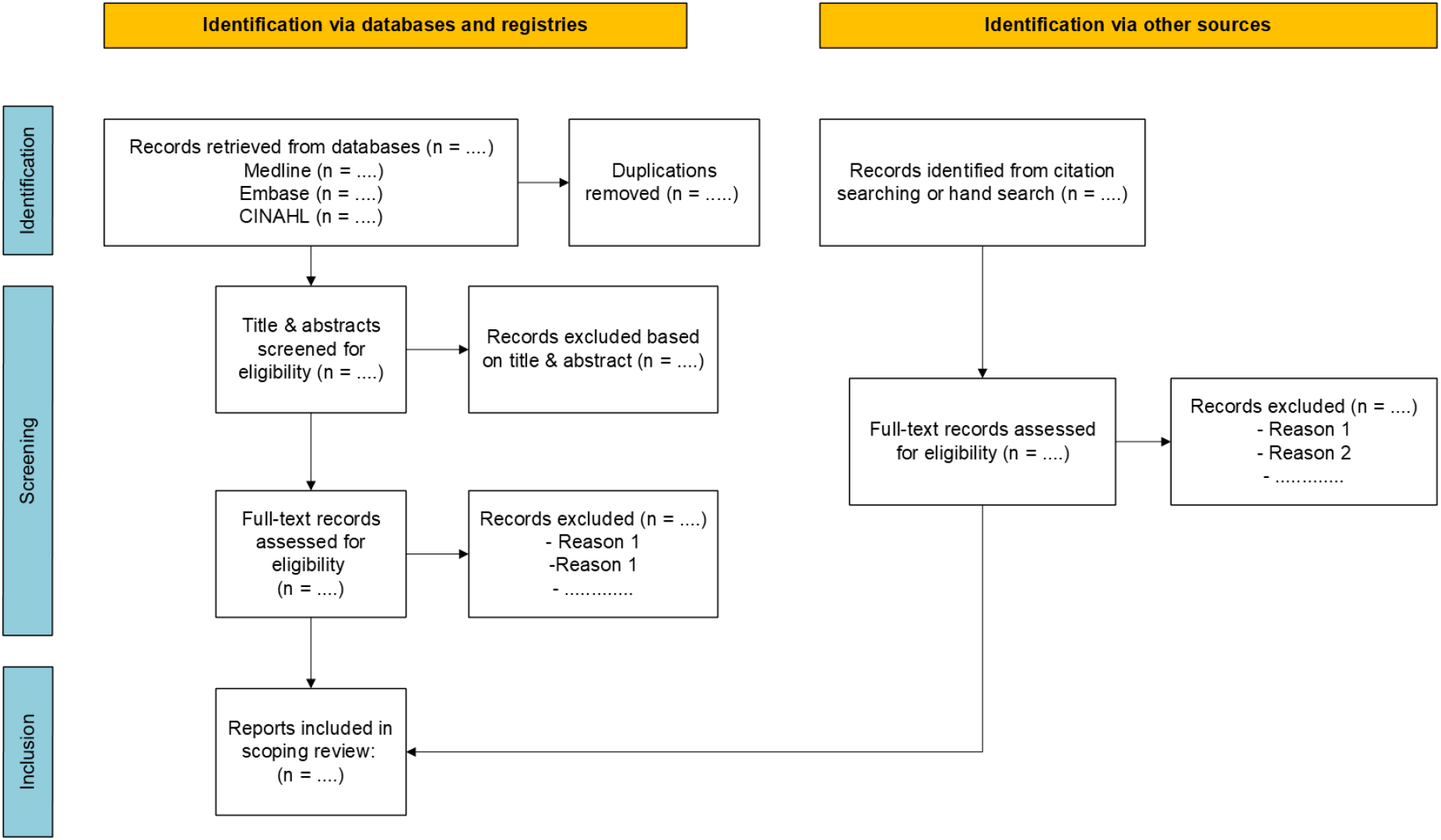
Flowchart of study selection.

### 1.4 Data Charting and Extraction

A draft version of the charting document can be found attached to this protocol (Supplemental 3). A data extraction form will be developed in Covidence and piloted with 10 publications by the authors; the data extraction form will contain the same data points as described in Supplemental 3:

a. Study background information: title, author(s), year of publication and country of origin, study design, study aim, population, sample size
b. Description of the form/mode of neonatal transport (ambulance, helicopter, etc.)
c. Description of the indication(s) for neonatal transport
  a. If applicable: intervention/exposure and comparator
d. Study methods
e. Information regarding the patient population (gestational age, birthweight, post-menstrual age at transport)
f. Outcome measures:
  a. If applicable: any information of the clinical situation/stability of the child before, during and/or after transport
  b. If applicable: any parent reported outcome and experience measures regarding transfer experiences
g. Key findings

Data will be extracted by one of the authors and will be verified by at least one other author. Any conflicts will be discussed with five authors (LAD, MLJP, MRK, AJA, JHLW) present. If the data cannot be extracted, or any additional information concerning a specific publication is needed, the corresponding author of the publication will be contacted as appropriate. Extracted data will be exported in XSLX format and used for analysis by the authors.

### 1.5 Quality assessment

Assessment of methodological quality is not typically part of a scoping review, however; due to the broad and explorative nature of this review, we aim to report on risk of bias in some capacity using the Mixed Methods Appraisal Tool (MMAT) by at least two authors in Covidence. No publications will be excluded based on this quality assessment.

### 1.6 Synthesis and Presentation of Results

Study selection will be presented in a PRISMA flowchart (see also: Figure 1). Background characteristics of the included publications will be presented in a descriptive table, which will include title, author(s), year of publication, country of origin, study design, population, and sample size, and appropriate referencing. If deemed useful, a visualization of the distribution of certain background characteristics (e.g. publication by year, country of origin or study design) will be added using Microsoft Excel (2016). Qualitative and quantitative outcomes will be reported separately. Further characterization will depend on the selection of articles that will eventually be included. For example, quantitative (health) outcomes could be further characterized as either short-term outcomes or long-term outcomes, or could be grouped by organ (heart, lungs, brain etc.).

### 1.7 Ethics and Dissemination

As this review is solely based on literature, and has no interventional or experimental features, an ethics review by the institutional review board is not necessary. This scoping review is intended to be published in a medical journal, as well as to be incorporated in the PhD thesis of one of the authors (LAD).

## Data Availability

All data produced in the present study are available upon reasonable request to the authors.

## Acknowledgements

The authors want to thank Dr. Wichor Bramer of the Erasmus MC Medical Library for his assistance with the creation of the search strategy for this scoping review.

