## Supplemental 1 for "Exploring the Impact of Non-Emergency Inter-Hospital Neonatal Transport of Preterm Infants: Study Protocol for a Scoping Review"

**Medline**

(Infant, Newborn/ OR exp Infant, Low Birth Weight/ OR Intensive Care Units, Neonatal/ OR Intensive Care, Neonatal/ OR Obstetric Labor, Premature/ OR (newborn* OR new*-born* OR neonat* OR nicu OR nicus OR prematur* OR preterm* OR pre-term* OR low-birth-weight OR lbw OR elbw OR vlbw).ab,ti,kw.) AND (exp Ambulances / OR (((patient* OR interhospital* OR inter-hospital* OR interinstit* OR inter-instit* OR neonat* OR new*-born* OR newborn*) ADJ3 (transport* OR transfer* OR referr* )) OR ambulance* OR helicopter*).ab,ti,kw. OR ((patient* OR interhospital* OR inter-hospital* OR interinstit* OR inter-instit* OR neonat* OR new*-born* OR newborn*) AND (transport* OR transfer* OR patient-referr*)).ti.) AND (exp Ambulances/ OR (interhospital* OR inter-hospital* OR interinstit* OR inter-instit* OR interfacilit* OR inter-facilit* OR internicu OR inter-nicu OR (between* ADJ3 (hospital* OR institute*)) OR ambulance* OR regionali*).ab,ti,kw. OR (hospital* OR institute* OR clinic OR clinics OR ((neonat* OR new*-born* OR newborn* OR preterm* OR pre-term*) ADJ6 (transport* OR transfer* OR referr*))).ti.) NOT (exp animals/ NOT humans/) AND (english.la. OR dutch.la.) AND (exp * Ambulances/ OR (transfer* OR transport* OR referr* OR ambulance* OR helicopter*).ti.)

**Embase**

(newborn/de OR 'low birth weight'/exp OR 'newborn care'/de OR 'newborn intensive care'/de OR 'neonatal intensive care unit'/de OR prematurity/de OR 'premature labor'/de OR (newborn* OR new*-born* OR neonat* OR nicu OR nicus OR prematur* OR preterm* OR pre-term* OR low-birth-weight OR lbw OR elbw OR vlbw):ab,ti,kw) AND ('patient transport'/exp OR 'patient referral'/de OR ambulance/exp OR 'air medical transport'/exp OR (((patient* OR interhospital* OR inter-hospital* OR interinstit* OR inter-instit* OR neonat* OR new*-born* OR newborn*) NEAR/3 (transport* OR transfer* OR referr* )) OR ambulance* OR helicopter*):ab,ti,kw OR ((patient* OR interhospital* OR inter-hospital* OR interinstit* OR inter-instit* OR neonat* OR new*-born* OR newborn*) AND (transport* OR transfer* OR patient-referr*)):ti) AND (ambulance/exp OR 'interhospital transfer'/de OR regionalization/de OR (interhospital* OR inter-hospital* OR interinstit* OR inter-instit* OR interfacilit* OR inter-facilit* OR internicu OR inter-nicu OR (between* NEAR/3 (hospital* OR institute*)) OR ambulance* OR regionali*):ab,ti,kw OR (hospital* OR institute* OR clinic OR clinics OR ((neonat* OR new*-born* OR newborn* OR preterm* OR pre-term*) NEAR/6 (transport* OR transfer* OR referr*))):ti) NOT [conference abstract]/lim NOT ([animals]/lim NOT [humans]/lim) AND ([english]/lim OR [dutch]/lim) AND ('patient transport'/exp/mj OR 'patient referral'/mj OR ambulance/exp/mj OR 'air medical transport'/exp/mj OR (transfer* OR transport* OR referr* OR ambulance* OR helicopter*):ti)

**CINAHL**

(MH Infant, Newborn OR MH Infant, Low Birth Weight+ OR MH Intensive Care Units, Neonatal OR MH Intensive Care, Neonatal OR MH Labor, Premature OR TI(newborn* OR new*-born* OR neonat* OR nicu OR nicus OR prematur* OR preterm* OR pre-term* OR low-birth-weight OR lbw OR elbw OR vlbw) OR AB(newborn* OR new*-born* OR neonat* OR nicu OR nicus OR prematur* OR preterm* OR pre-term* OR low-birth-weight OR lbw OR elbw OR vlbw)) AND (MH Ambulances + OR TI(((patient* OR interhospital* OR inter-hospital* OR interinstit* OR inter-instit* OR neonat* OR new*-born* OR newborn*) N2 (transport* OR transfer* OR referr* )) OR ambulance* OR helicopter*) OR AB(((patient* OR interhospital* OR inter-hospital* OR interinstit* OR inter-instit* OR neonat* OR new*-born* OR newborn*) N2 (transport* OR transfer* OR referr* )) OR ambulance* OR helicopter*) OR TI((patient* OR interhospital* OR inter-hospital* OR interinstit* OR inter-instit* OR neonat* OR new*-born* OR newborn*) AND (transport* OR transfer* OR patient-referr*))) AND (MH Ambulances+ OR TI(interhospital* OR inter-hospital* OR interinstit* OR inter-instit* OR interfacilit* OR inter-facilit* OR internicu OR inter-nicu OR  (between* N2 (hospital* OR institute*)) OR ambulance* OR regionali*) OR AB(interhospital* OR inter-hospital* OR interinstit* OR inter-instit* OR interfacilit* OR inter-facilit* OR internicu OR inter-nicu OR  (between* N2 (hospital* OR institute*)) OR ambulance* OR regionali*) OR TI(hospital* OR institute* OR clinic OR clinics OR ((neonat* OR new*-born* OR newborn* OR preterm* OR pre-term*) N5 (transport* OR transfer* OR referr*)))) NOT (MH animals+ NOT MHhumans+) AND LA(english OR dutch) AND (MM Ambulances+ OR TI(transfer* OR transport* OR referr* OR ambulance* OR helicopter*))
