## Supplemental 3 for "Exploring the Impact of Non-Emergency Inter-Hospital Neonatal Transport of Preterm Infants: Study Protocol for a Scoping Review"

| Title | Author(s) | Year of publication | Country of origin | Study aim | Study design | Population | Sample size | Description of form/mode of neonatal transport | Description of indication(s) for neonatal transport | If applicable: type of intervention or exposure, and if present the comparator | Description of methods | Patient population: GA | Patient population: BW | Patient population: PMA transport | Description of outcome measures | If applicable: clinical situation pre-transport | If applicable: clinical situation during transport | If applicable: clinical situation post-transport | Key findings |
|---|---|---|---|---|---|---|---|---|---|---|---|---|---|---|---|---|---|---|---|

Abbreviations: GA, gestational age; BW, birthweight; PMA, post-menstrual age.
